# Are the relevant risk factors being adequately captured in empirical studies of smoking initiation? A machine learning analysis based on the Population Assessment of Tobacco and Health study

**DOI:** 10.1101/2022.09.18.22280076

**Authors:** Thuy T. T. Le, Mona Issabakhsh, Yameng Li, Luz María Sánchez-Romero, Jiale Tan, Rafael Meza, David Levy, David Mendez

**Affiliations:** University of Michigan, School of Public Health, Department of Health Management and Policy, Ann Arbor, MI, USA; University of Michigan School of Public Health, Department of Epidemiology, Ann Arbor, MI, USA; Georgetown University-Lombardi Comprehensive Cancer Center, Washington, DC, USA; Integrative Oncology, BC Cancer Research Institute, Vancouver BC

## Abstract

Cigarette smoking continues to pose a threat to public health. Identifying individual risk factors for smoking initiation is essential to further mitigate this epidemic. To our knowledge, no study today has used Machine Learning (ML) techniques to automatically uncover informative predictors of smoking onset among adults using the Population Assessment of Tobacco and Health (PATH) study. In this work, we employed Random Forest paired with Recursive Feature Elimination to identify relevant PATH variables that predict smoking initiation among adult never smokers at baseline between two consecutive PATH waves. We included all potentially informative baseline variables in wave 1 (wave 4) to predict past 30-day smoking status in wave 2 (wave 5). Using the first and most recent pairs of PATH waves was found sufficient to identify the key risk factors of smoking initiation and test their robustness over time. As a result, classification models suggested about 60 informative PATH variables among more than 200 candidate variables in each baseline wave. With these selected predictors, the resulting models have a high discriminatory power with the area under the Specificity-Sensitivity curves of around 80%. We examined the chosen variables and discovered important features. Across the considered waves, three factors, (i) BMI, (ii) dental/oral health status, and (iii) taking anti-inflammatory or pain medication, robustly appeared as significant predictors of smoking initiation, besides other well-established predictors. Our work demonstrates that ML methods are useful to predict smoking initiation with high accuracy, identify novel smoking initiation predictors, and enhance our understanding of tobacco use behaviors.

## Introduction

During the past decade, the tobacco product landscape has evolved rapidly with a remarkable decrease in cigarette smoking prevalence [1], a dramatic increase in the popularity of electronic nicotine delivery systems (ENDS), and the emergence of other novel tobacco products such as heated tobacco products [2, 3]. However, cigarette smoking is still the main cause of tobacco-related morbidity and mortality, being responsible for hundreds of thousands of premature deaths and costing the United States (US) economy hundreds of billions of dollars annually [4, 5]. Despite the significant decline in smoking prevalence, reducing initiation rates and increasing cessation rates remain the focus of tobacco control to further reduce the burden of tobacco use and prevent a potential reversal in the reductions of population-level smoking rates from the past decades. To better support tobacco regulations, more studies predicting smoking behaviors as well as identifying risk factors of such behaviors are needed.

Using survey data and traditional statistical models such as logistic regression, previous research has uncovered a list of individual-specific characteristics associated with smoking initiation. Due to the standard assumptions of these models and the correlation between potential explanatory variables, these analyses can generally only incorporate a relatively small number of expert-selected predictors [6-9]. Consequently, they generally do not exploit the vast amount of information in survey data such as the many variables collected by the US Population Assessment of Tobacco and Health (PATH) study [10]. In addition, the findings from traditional statistical models are likely to depend on the considered risk factors, which may lead to different conclusions if other factors are considered [6] and the omission of other unanticipated risk factors. Furthermore, previous studies have shown that these traditional methods may not perform optimally as prediction models, especially when dealing with class-imbalanced data (having an imbalance between class samples) [11, 12]. Therefore, in predicting individuals’ smoking behaviors, alternative methods are needed.

Machine learning (ML) methods such as random forest (RF) are powerful tools that can address some of these limitations. Many ML algorithms can automatically identify hidden patterns in complex data that researchers would otherwise struggle to uncover to predict future outcomes. Applications of machine learning techniques have just started infiltrating tobacco control research, despite the popularity of its applications in other research fields [13]. In tobacco control, a growing number of research articles have focused on predicting smoking and vaping behaviors and identifying the most significant predictors of such behaviors through ML algorithms. For instance, researchers used ML methods such as RF classifiers and penalized logistic regression models to predict ENDS use status and identify the most important predictors of this behavior based on various surveys [13-15]. Coughlin et. al. employed decision trees to classify smoking cessation outcomes in group cognitive-behavioral therapy at a 6-month follow-up and identify the most relevant risk factors of treatment outcomes [16]. Kim and colleagues applied regression tree models to identify complex interactions among various factors that differentiate global adolescents’ levels of tobacco susceptibility and use [17].

Most studies used data from clinical trials or local representative surveys [13, 15, 16, 18]. Therefore, the findings may not be generalizable to the whole US population. The PATH study has been a primary data source of research work on population-level tobacco use behaviors [10]. PATH is a large, longitudinal, and nationally representative survey [10]. It contains detailed individual-specific information, including demographics, socioeconomic and health status, perceptions, substance use, and tobacco use. To our knowledge, no study to date has applied ML models to predict smoking initiation and identify relevant predictors of this behavior in the adult PATH data. Here we employed an RF classifier paired with Recursive Feature Elimination (RFE), popular ML methods, to predict smoking initiation among adult never smokers at baseline between two consecutive PATH waves and identify critical baseline factors associated with smoking initiation. Our analysis incorporates all PATH variables relevant to the smoking onset. This study is intended to enrich the literature on applications of ML methods in tobacco control, advance our understanding of smoking initiation, and aid tobacco regulators in designing targeted tobacco interventions.

## Methods

### Data preparation

The PATH survey is a national longitudinal survey of tobacco use and its effects on the US population. PATH conducted its first wave in 2013 and has publicly released 5 waves of adult data (wave 1: September 2013–December 2014, wave 2: October 2014–October 2015, wave 3: October 2015–October 2016, wave 4: December 2016—January 2018, and wave 5: December 2018–November 2019) [10]. Using the PATH survey, we aimed to predict smoking initiation between two consecutive waves among adult never smokers at baseline and identify relevant predictors of this behavior. Here, individuals who never tried smoking even one or two puffs of a cigarette are referred to as never smokers. In PATH, a nationally representative sample of adults was surveyed in wave 1, completing follow-up interviews at each follow-up wave. In wave 4, a new replenishment sample was added to the study to address attrition over time and reset the sample size, making it again nationally representative.

In this study, we analyzed the earliest (waves 1 and 2) and more recent (waves 4 and 5) pairs of waves. Considering these two pairs of waves we identified all key risk factors of smoking initiation in PATH and investigated whether the key factors for smoking initiation changed over time. We used the wave 1 (wave 4) variables to predict the past 30-day (P30D) smoking status in wave 2 (wave 5). The P30D smoking status of an individual is defined as ‘Yes’ if he/she smoked a cigarette in the past 30 days and ‘No’ otherwise.

For waves 1 and 2, we first extracted a subpopulation of never smokers in wave 1 and combined it with the outcome variable that indicates the P30D smoking status in wave 2 using individuals’ identity numbers. We then removed non-relevant variables such as personal identity numbers, random questions, sample weights, and imputed variables. Furthermore, variables with more than 5% missing values of the total sample size were dropped to maintain the highest possible number of predictors. We also excluded individuals with missing smoking status in wave 2. Finally, to use these datasets for model development later, all the remaining missing values were imputed using a supervised imputation method [19]. The final analytic dataset included data from 5776 adults with 209 predictors, see Table 1.

**Table 1:** A description of the clean and complete datasets extracted from the PATH data after data processing.

| Baseline smoking status | Outcome smoking status | Value of outcome smoking status |  | Number of predictors | Total |
| --- | --- | --- | --- | --- | --- |
|  |  | Yes | No |  |  |
| Never smokers in wave 1 | P30D smoking status in wave 2 | 197<br>(3.4%) | 5579<br>(96.6%) | 209 | 5776<br>(100%) |
| Never smokers in wave 4 | P30D smoking status in wave 5 | 208<br>(2.6%) | 7687<br>(97.4%) | 293 | 7895<br>(100%) |

However, as shown in Table 1, most wave 1 never smokers (more than 95%) remained as never smokers in wave 2. Therefore, the distribution of the outcome P30D smoking status in wave 2 is highly imbalanced, which often has a negative impact on predicting the minority class (i.e., P30D smokers). To overcome this issue and improve the classification power of the model, we divided each dataset into training and test data (60%:40%) and created balanced training data by randomly oversampling minority examples [20], while the test data remained untouched. The balanced training data was used to train and validate the chosen model classifier.

Analogously, we repeated this whole process and obtained the final clean dataset for waves 4 and 5 (7895 adults with 293 predictors) shown in Table 1. A difference of 84 predictors between these two pairs of waves (209 predictors for waves 1 and 2 versus 293 predictors for waves 4 and 5) is due to the changes in the PATH survey questionnaires between Wave 1 and Wave 4.

### Statistical analysis

After data preprocessing, we obtained two datasets corresponding to two pairs of waves in Table 1. Our datasets with more than 200 variables are likely to contain many redundant or irrelevant predictors. Searching for an optimal subset of predictors is usually needed to produce more effective data mining algorithms, improve the predictive accuracy of the considered model, and increase the comprehensibility of the underlying process that generated the data [21]. As such, for each pair of waves, we used an RF classifier combined with RFE (RF-RFE) to obtain a subset of predictors on which the RF classifier performs best [22]. RF is an ensemble method that consists of many decision trees trained in parallel on different subsets bootstrapped from the original data. The final RF decision is generated by aggregating the decisions of all individual decision trees. For RF-RFE, the RF classifier was, first, trained on the entire training set, providing a rank order of all the predictors based on the mean decrease of accuracy. The mean decrease in accuracy was computed by randomly permuting the values of each predictor. Variables whose permutation causes a significant loss in accuracy correspond with high importance scores. Variable(s) with the lowest important ranking was (were) then removed from the dataset. The model was re-trained on the reduced predictors, and its performance was recorded. This process was repeated until all the predictors were explored [22]. We trained RF-RFE on the balanced training data with 5-fold cross-validation repeated 3 times to score different subsets of predictors, select the best subset of predictors, and avoid selection bias [23]. The best predictor subset may contain all predictors if they are all informative. The test set was used to evaluate how well the model performance would generalize to new data drawn from the same distribution but not included in the training data. For more details on how RF-RFE works, we refer the reader to Kuhn’s work [23]. Here, we chose RF as a core method for RFE due to its appealing features including high prediction accuracy, ability to deal with high dimensional data, and internal measure of variable importance [24, 25].

### Performance measures of the RF classifier

A receiver operating characteristic (ROC) curve is a graphical representation of a classifier’s performance as a tradeoff between Specificity and Sensitivity. It is typically obtained by plotting the true positive rate (Sensitivity) against the true negative rate (Specificity) for a single classifier at various probability cutoff values. For each outcome wave,

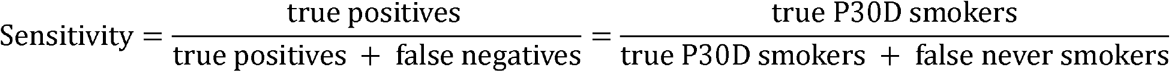

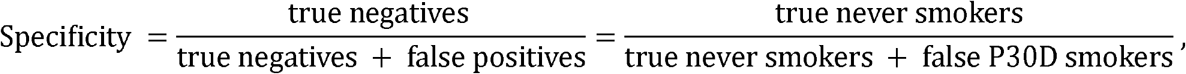

where true P30D smokers (false never smokers) are the actual P30D smokers in the outcome wave who are correctly (incorrectly) predicted by the model, and true never smokers (false P30D smokers) are the actual never smokers in the outcome wave who are correctly (incorrectly) predicted by the model. Sensitivity and Specificity show the ability of a classifier to correctly identify P30D smokers in wave 2 (wave 5) among wave 1 (wave 4) never smokers. The area under the ROC curve (AUC) is usually a preferred measure of the model performance in binary classification problems. AUC is a more discriminating measure of performance than accuracy in classification [26]. The closer AUC gets to 100%, the better the classifier performance. A perfect classifier corresponds to an AUC of 100%, while a classifier without any predictive power corresponds to an AUC of 50%. Finally, to provide a reliable estimate of AUC, we also computed the 95% confidence interval (CI) of AUC with 2000 stratified bootstrap replicates.

The study was exempted from the review of the Institutional Review Board at the authors’ institution because of the public availability of the data.

## Results

### Most relevant predictors of smoking initiation

RF-RFE selected a list of only about 60 relevant variables of the 200+ possible variables (64 and 60 in waves 1 and 4, respectively) for predicting smoking initiation among adult never smokers at baseline as shown in detail in the Appendix. With these recommended variables, the classification model’s performance metrics (Accuracy and Kappa [27]) reach their best levels. The sets of selected variables from the baseline waves 1 and 4 contain many highly correlated predictors, see the Appendix. Thus, the rankings of predictors produced by the RF classifier may be biased due to variable correlations [28, 29]. Focusing on justifying only a few top RF-ranked variables as the most important predictors of the interesting outcomes like in previous studies [14, 15, 18] is unreliable. Therefore, we considered all variables relevant to the outcome variables. Based on the description of each variable, we grouped them into different categories as detailed in the Appendix. Variables describing the same characteristics would be put into the same category.

As a result, we obtained a set of individual characteristics associated with the onset of cigarette smoking, see Table 2. Table 2 indicates that, across the considered baseline waves, age, BMI, dental/oral care, education, employment status, experimentation with other tobacco products, exposure to radio/TV, financial status, mental/physical health status, race, social influence, taking anti-inflammatory or pain medication, tobacco risk perceptions/awareness are important factors associated with smoking initiation by the following wave. In addition to those variables, exposure to tobacco products/tobacco advertisements, gender/sexual orientation, household rules about tobacco use, family size, and internet access are related to smoking initiation between waves 1 and 2. However, exposure to anti-tobacco advertisements, marital status, and smoking intention were predictive factors identified only in the wave 4 to 5 analysis.

**Table 2:** Individuals’ characteristics related to the transition from never smokers to new smokers between two consecutive waves.

| Wave 1 | Wave 4 |
| --- | --- |
| Age<br>BMI<br>Dental/Oral care<br>Education<br>Employment status<br>Experimentation with other tobacco products<br>Exposure to radio/TV<br>Financial status<br>Mental health status<br>Physical health status<br>Race<br>Social influence<br>Taking anti-inflammatory or pain medication<br>Tobacco risk perceptions/awareness |  |
| Exposure to tobacco products/ tobacco advertisements<br>Gender/Sexual orientation<br>Household rules about tobacco use<br>Household size<br>Internet use | Exposure to anti-tobacco advertisements<br>Smoking intention<br>Marital status |

Lists of the top 20 relevant predictors with their rankings are presented in the Appendix.

### Performance of the RF classifier

Table 3 shows the predictive power of the trained RF classifiers on the test data using the selected variables (around 60 variables). Critical metrics for evaluating the classifier’s performance including AUCs, Specificity, and Sensitivity with their 95% CIs are computed. Column 3 indicates that the RF classifier performs very well on all considered datasets with AUCs of around 80%. Column 3 displays the optimal probability cutoffs which are the closest points to the top-left parts of the plots with perfect Sensitivity and Specificity. These thresholds are computed using the *closest-topleft* method [30].

**Table 3:** The key performance metrics of the RF classifiers.

|  | AUC<br>(95% CI) | Optimal<br>threshold | Specificity<br>(95% CI) | Sensitivity<br>(95% CI) |
| --- | --- | --- | --- | --- |
| Waves 1 and 2 | 80.1%<br>(75.4%-84.9%) | 0.949 | 78.2%<br>(69.2%-87.2%) | 70.9%<br>(69.0%-72.7%) |
| Waves 4 and 5 | 79.6%<br>(75.0%-84.2%) | 0.959 | 75.9%<br>(66.3%-84.3%) | 69.5%<br>(67.9%-71.1%) |

Individuals with predicted probability greater than these thresholds are classified as never smokers and P30D smokers otherwise. Columns 4 and 5 are the values of Sensitivity and Specificity corresponding with the optimal thresholds in Column 3. Increasing Sensitivity, more P30D smokers identified correctly, leads to a decrease in Specificity (more never smokers would be incorrectly identified) and vice versa. Therefore, these thresholds are just suggestive and should be adjusted to achieve one’s desirable balance between Sensitivity and Specificity. The ROC curves of the RF classifiers are plotted in Figure 1.

**Figure 1:**
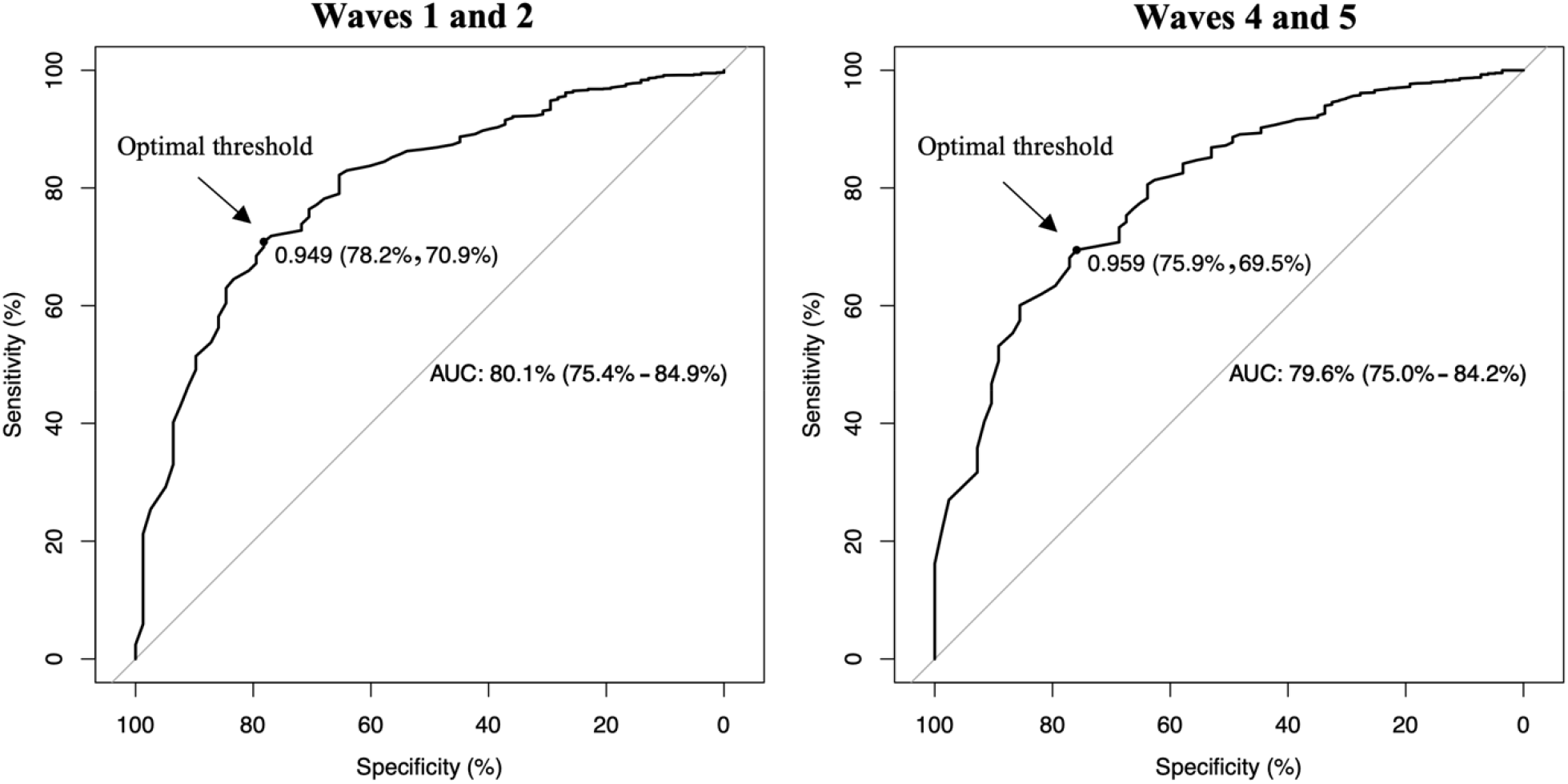
The ROC curves of all the RF classifiers. The diagonal line in each plot represents random guessing. Each plot shows the AUC curve (95% CI) together with the optimal threshold (Specificity, Sensitivity).

## Discussion

This is the first study to leverage all possible original public adult PATH variables to predict the transition from never to P30D cigarette smokers between two consecutive waves using RF-RFE. Despite the severely skewed class distribution shown in Table 1, the model performs well in classifying smoking status among never smokers with AUC = 80% approximately (see Table 3 and Figure 1). We uncovered about 60 variables associated with the smoking onset between two considered PATH waves among adult never smokers at baseline. The findings in Table 2 show that most of the discovered risk factors are robust across waves and their association with smoking initiation is well-reported in the literature.

For the joint predictive variables between waves 1 and 4, most of them (i.e., age [31-34], education [33, 34], employment status [35], experimentation with other tobacco products [34, 36-40], exposure to radio/TV [41], financial status [34, 42], mental/physical health status [43], race [33, 34, 44], social influence [45-47], and tobacco risk perceptions/awareness [33, 47]) are familiar risk factors of smoking initiation. Meanwhile, other variables including BMI, dental/oral health status, and taking anti-inflammatory or pain medication, have not been consistently identified as initiation predictors in the literature. Across the considered waves, our model consistently indicates BMI as one of the main predictors of the onset of cigarette smoking in the following waves. Several studies have found a strong connection between BMI and smoking initiation while analyzing different datasets [47-50]. When RF-RFE was applied to these datasets stratified by gender, BMI remained among the relevant predictors regardless of gender. This association could be related to weight concerns since many believe that smoking helps control their weight or via other sociodemographic factors such as lower socioeconomic status, income, and educational attainment, which are highly correlated to BMI [47-50]. Oral/dental care robustly appears among the predictors of smoking initiation in both baseline waves. The association between oral/dental care and smoking onset has not been widely reported in the literature, except in earlier studies that analyzed data from a randomized clinical trial in California to predict tobacco initiation over a two-year follow-up interval among adolescent never tobacco users [50, 51]. People who take good care of their oral health may be less likely to start smoking since they are probably aware of the harmful impacts of smoking on their teeth and gum health. Furthermore, our findings indicate that taking anti-inflammatory or pain medication is also associated with smoking initiation. Individuals taking these medications are likely to be in bad physical/mental health status and therefore find smoking a way of stress/pain relief [52]. There could be other possible explanations for these connections. More work is required to investigate and understand the impacts of these factors on smoking initiation.

Besides the shared predictors of smoking initiation across the considered waves, each baseline wave has its own set of relevant predictors, in part due to the changes in the PATH questionnaire. Again, most wave-specific variables are well reported in previous studies except internet use in wave 1. This variable is likely to indicate exposure to smoking-stimulating factors, including onscreen tobacco use, tobacco advertisements, and digitally social influence. A recent study [14] reported a similar observation while using a penalized logistic regression model on the PATH data to predict the use of ENDS among never users of any tobacco products. These authors found an association between digital media use with future ENDS use when using different waves in U.S. adolescents.

Like other similar studies [14, 18], our work has limitations. First, since the PATH survey is self-reported, the responses are potentially subject to recall bias. Second, our analysis was conducted based on the unweighted PATH data, therefore our results may not generalize to the whole population.

However, since the PATH survey is nationally representative, i.e., the survey sample captures the characteristics of most US individuals, these conclusions are likely to hold in general. However, external validation of the resulting models is needed. Third, factors not included in the PATH study were not considered. Fourth, the list of all relevant factors of smoking onset was recommended by RF-RFE. Other ML methods may provide a slightly different set of these factors. Finally, it should be noted that our results cannot establish the causal direction of the association of the model-selected predictors with smoking initiation due to the nature of the RF algorithm.

In summary, this work demonstrates the potential applications of machine learning methods in developing highly accurate classification models to predict smoking initiation using national representative survey data and systematically identify all potential risk factors of smoking behaviors in the PATH data. Besides reconfirming well-known risk factors in the literature, our findings discovered additional predictors of smoking initiation that have not been paid much attention to in previous studies. More studies that focus on the newly discovered factors are needed to confirm their predictive power against changes in smoking behaviors as well as determine the underlying mechanisms.

## Data Availability

All data produced are available online at https://www.icpsr.umich.edu/web/NAHDAP/studies/36231

## Data availability

All data produced are available online at https://www.icpsr.umich.edu/web/NAHDAP/studies/36231

## CRediT authorship contribution statement

**Thuy T. T. Le**: Conceptualization, Methodology, Formal Analysis, Software, Data Curation, Writing - Original Draft, Visualization, Funding Acquisition. **Mona Issabakhsh**: Conceptualization, Writing - Review & Editing, Funding Acquisition. **Yameng Li, Luz María Sánchez-Romero** and **Jiale Tan**: Writing - Review & Editing. **Rafael Meza, David Levy** and **David Mendez**: Conceptualization, Writing - review & editing, Supervision.

## References

[1] Cornelius ME, Wang TW, Jamal A, et al. Tobacco product use among adults—United States, 2019. Morbidity and Mortality Weekly Report 2020;69(46):1736.

[2] US Department of Health and Human Service. Surgeon General’s advisory on e-cigarette use among youth. https://e-cigarettes.surgeongeneral.gov/documents/surgeon-generals-advisory-on-e-cigarette-use-among-youth-2018.pdf. Accessed May 3rd 2022.

[3] US Department of Health and Human Services. E-cigarette use among youth and young adults: A report of the Surgeon General. 2016.

[4] U.S. Department of Health and Human Services. The health consequences of smoking - 50 years of progress: a report of the Surgeon General. Atlanta, GA: U.S. Department of Health and Human Services, Centers for Disease Control and Prevention, Office on Smoking and Health 2014.

[5] Xu X, Shrestha SS, Trivers KF, et al. US healthcare spending attributable to cigarette smoking in 2014. Preventive Medicine 2021;150:106529.

[6] Sun R, Mendez D, Warner KE. Is Adolescent E-Cigarette Use Associated With Subsequent Smoking? A New Look. Nicotine & Tobacco Research 2021;24(5):710–718.

[7] Loukas A, Marti CN, Cooper M, et al. Exclusive e-cigarette use predicts cigarette initiation among college students. Addictive behaviors 2018;76:343–347.

[8] Watkins SL, Glantz SA, Chaffee BW. Association of noncigarette tobacco product use with future cigarette smoking among youth in the Population Assessment of Tobacco and Health (PATH) study, 2013-2015. JAMA pediatrics 2018;172(2):181–187.

[9] Bell K, Keane H. All gates lead to smoking: the ‘gateway theory’, e-cigarettes and the remaking of nicotine. Social Science & Medicine 2014;119:45–52.

[10] FDA and NIH Study: Population Assessment of Tobacco and Health. https://www.fda.gov/tobacco-products/research/fda-and-nih-study-population-assessment-tobacco-and-health. Accessed July 14th 2022.

[11] Couronné R, Probst P, Boulesteix A-L. Random forest versus logistic regression: a large-scale benchmark experiment. BMC bioinformatics 2018;19(1):1–14.

[12] Muchlinski D, Siroky D, He J, et al. Comparing random forest with logistic regression for predicting class-imbalanced civil war onset data. Political Analysis 2016;24(1):87–103.

[13] Fu R, Kundu A, Mitsakakis N, et al. Machine learning applications in tobacco research: a scoping review. Tobacco Control 2021.

[14] Han D-H, Lee SH, Lee S, et al. Identifying emerging predictors for adolescent electronic nicotine delivery systems use: A machine learning analysis of the Population Assessment of Tobacco and Health Study. Preventive Medicine 2021;145:106418.

[15] Shi J, Fu R, Hamilton H, et al. A machine learning approach to predict e-cigarette use and dependence among Ontario youth. Health Promotion and Chronic Disease Prevention in Canada: Research, Policy and Practice 2022;42(1):21.

[16] Coughlin LN, Tegge AN, Sheffer CE, et al. A machine-learning approach to predicting smoking cessation treatment outcomes. Nicotine and Tobacco Research 2020;22(3):415–422.

[17] Kim N, Loh W-Y, McCarthy DE. Machine learning models of tobacco susceptibility and current use among adolescents from 97 countries in the Global Youth Tobacco Survey, 2013-2017. PLOS Global Public Health 2021;1(12):e0000060.

[18] Fu R, Shi J, Chaiton M, et al. A Machine Learning Approach to Identify Predictors of Frequent Vaping and Vulnerable Californian Youth Subgroups. Nicotine and Tobacco Research 2022;24(7):1028–1036.

[19] RColor Brewer S, Liaw MA. Package ‘randomforest’. University of California, Berkeley: Berkeley, CA, USA 2018.

[20] Lunardon N, Menardi G, Torelli N, et al. Package ‘ROSE’. 2021.

[21] Saeys Y, Inza I, Larranaga P. A review of feature selection techniques in bioinformatics. bioinformatics 2007;23(19):2507–2517.

[22] Guyon I, Weston J, Barnhill S, et al. Gene selection for cancer classification using support vector machines. Machine learning 2002;46(1):389–422.

[23] Kuhn M. Variable selection using the caret package. URL http://crancerminlipigoid/web/packages/caret/vignettes/caretSelectionpdf 2012:1–24.

[24] Biau G, Scornet E. A random forest guided tour. Test 2016;25(2):197–227.

[25] Cutler A, Cutler DR, Stevens JR. Random forests. Ensemble machine learning: Springer 2012:157–175.

[26] Ling CX, Huang J, Zhang H. AUC: a statistically consistent and more discriminating measure than accuracy. Ijcai 2003:519–524.

[27] McHugh ML. Interrater reliability: the kappa statistic. Biochemia medica 2012;22(3):276–282.

[28] Toloşi L, Lengauer T. Classification with correlated features: unreliability of feature ranking and solutions. Bioinformatics 2011;27(14):1986–1994.

[29] Strobl C, Boulesteix A-L, Zeileis A, et al. Bias in random forest variable importance measures: Illustrations, sources and a solution. BMC bioinformatics 2007;8(1):1–21.

[30] Robin X, Turck N, Hainard A, et al. Package ‘pROC’. Package ‘pROC’ 2021.

[31] Barrington-Trimis JL, Braymiller JL, Unger JB, et al. Trends in the age of cigarette smoking initiation among young adults in the US from 2002 to 2018. JAMA network open 2020;3(10):e2019022–e2019022.

[32] Escobedo LG, Anda RF, Smith PF, et al. Sociodemographic characteristics of cigarette smoking initiation in the United States: implications for smoking prevention policy. Jama 1990;264(12):1550–1555.

[33] Freedman KS, Nelson NM, Feldman LL. Smoking initiation among young adults in the United States and Canada, 1998-2010: a systematic review. Preventing chronic disease 2012;9.

[34] Kasza KA, Edwards KC, Tang Z, et al. Correlates of tobacco product initiation among youth and adults in the USA: findings from the PATH Study Waves 1–3 (2013–2016). Tobacco control 2020;29(Suppl 3):s191–s202.

[35] Marcus J. Does job loss make you smoke and gain weight? Economica 2014;81(324):626–648.

[36] Aleyan S, Cole A, Qian W, et al. Risky business: a longitudinal study examining cigarette smoking initiation among susceptible and non-susceptible e-cigarette users in Canada. BMJ open 2018;8(5):e021080.

[37] Leventhal AM, Strong DR, Kirkpatrick MG, et al. Association of electronic cigarette use with initiation of combustible tobacco product smoking in early adolescence. Jama 2015;314(7):700–707.

[38] Soneji S, Barrington-Trimis JL, Wills TA, et al. Association Between Initial Use of e-Cigarettes and Subsequent Cigarette Smoking Among Adolescents and Young Adults: A Systematic Review and Meta-analysis. JAMA pediatrics 2017;171(8):788–797.

[39] Baenziger ON, Ford L, Yazidjoglou A, et al. E-cigarette use and combustible tobacco cigarette smoking uptake among non-smokers, including relapse in former smokers: umbrella review, systematic review and meta-analysis. BMJ open 2021;11(3):e045603.

[40] Coreas SI, Rodriquez EJ, Rahman SG, et al. Smoking susceptibility and tobacco media engagement among youth never smokers. Pediatrics 2021;147(6).

[41] Johnson EK, Len-Ríos M, Shoenberger H, et al. A fatal attraction: The effect of TV viewing on smoking initiation among young women. Communication Research 2019;46(5):688–707.

[42] Binkley J. Low income and poor health choices: the example of smoking. American Journal of Agricultural Economics 2010;92(4):972–984.

[43] Rohde P, Lewinsohn PM, Brown RA, et al. Psychiatric disorders, familial factors and cigarette smoking: I. Associations with smoking initiation. Nicotine & Tobacco Research 2003;5(1):85–98.

[44] Thompson AB, Mowery PD, Tebes JK, et al. Time trends in smoking onset by sex and race/ethnicity among adolescents and young adults: findings from the 2006–2013 National Survey on Drug Use and Health. Nicotine and Tobacco Research 2018;20(3):312–320.

[45] Chezhian C, Murthy S, PraSad S, et al. Exploring factors that influence smoking initiation and cessation among current smokers. Journal of clinical and diagnostic research: JCDR 2015;9(5):LC08.

[46] Khalil GE, Jones EC, Fujimoto K. Examining proximity exposure in a social network as a mechanism driving peer influence of adolescent smoking. Addictive Behaviors 2021;117:106853.

[47] Bernat DH, Klein EG, Forster JL. Smoking initiation during young adulthood: a longitudinal study of a population-based cohort. Journal of Adolescent Health 2012;51(5):497–502.

[48] Taylor AE, Richmond RC, Palviainen T, et al. The effect of body mass index on smoking behaviour and nicotine metabolism: a Mendelian randomization study. Human molecular genetics 2019;28(8):1322–1330.

[49] Murphy CM, Janssen T, Colby SM, et al. Low self-esteem for physical appearance mediates the effect of body mass index on smoking initiation among adolescents. Journal of Pediatric Psychology 2019;44(2):197–207.

[50] Wahlgren DR, Hovell MF, Slymen DJ, et al. Predictors of tobacco use initiation in adolescents: a two-year prospective study and theoretical discussion. Tobacco Control 1997;6(2):95–103.

[51] Hovell MF, Slymen DJ, Keating KJ, et al. Tobacco use prevalence and correlates among adolescents in a clinician initiated tobacco prevention trial in California, USA. Journal of Epidemiology & Community Health 1996;50(3):340–346.

[52] Patterson AL, Gritzner S, Resnick MP, et al. Smoking cigarettes as a coping strategy for chronic pain is associated with greater pain intensity and poorer pain-related function. The Journal of Pain 2012;13(3):285–292.

